# Distinct Patterns of Pituitary Dysfunction in Combination Immunotherapy with Nivolumab and Ipilimumab

**DOI:** 10.1101/2025.05.16.25327742

**Authors:** Aya Sakurai, Takuro Hakata, Ichiro Yamauchi, Sadahito Kimura, Daisuke Kosugi, Taku Sugawa, Haruka Fujita, Kentaro Okamoto, Yohei Ueda, Daisuke Taura, Daisuke Yabe

## Abstract

**Context:** Immune checkpoint inhibitors (ICIs) have revolutionized cancer treatment but are associated with immune-related adverse events (irAEs), including pituitary dysfunction. Combination immunotherapy with PD-1 and CTLA-4 inhibitors increases the risk of pituitary irAEs compared to PD-1 monotherapy; however, their detailed clinical characteristics remain unclear.

**Objective:** To clarify the clinical features of pituitary irAEs induced by combination immunotherapy.

**Methods:** In this retrospective cohort study, we compared patients treated with combination therapy of nivolumab and ipilimumab (Nivo/Ipi) to those receiving nivolumab monotherapy (Nivo). We analyzed clinical data including presenting symptoms, laboratory findings, pituitary MRI results, and coexisting irAEs.

**Results:** Pituitary irAEs were more frequent in the Nivo/Ipi group (17.4%) than in the Nivo group (2.7%) and developed earlier (median onset: 63 vs. 153 days, respectively). All 15 patients in the Nivo group presented with isolated ACTH deficiency (IAD), whereas the Nivo/Ipi group included 8 cases of IAD and 8 cases of combined pituitary hormone deficiency (CPHD). In the Nivo/Ipi group, CPHD occurred significantly earlier than IAD (median onset: 40 vs. 84 days) and was associated with a higher incidence of headache and pituitary swelling on MRI. Furthermore, 75% of patients with CPHD also experienced non-endocrine irAEs. Two CPHD patients experienced worsening of symptoms and pituitary dysfunction following re-administration of Nivo/Ipi.

**Conclusion:** Pituitary irAEs are more frequent and develop earlier in patients receiving Nivo/Ipi. CPHD and IAD, induced by this combination immunotherapy, exhibit distinct clinical courses. Recognizing these differences is crucial for the optimal management of pituitary irAEs during combination immunotherapy.

## Introduction

In recent years, immune checkpoint inhibitors (ICIs) have become essential therapeutic agents in the treatment of various cancers. However, the use of ICIs is often associated with immune-related adverse events (irAEs), and their management is crucial for the success of immunotherapy (1,2). Monoclonal antibodies against cytotoxic T-lymphocyte–associated protein 4 (CTLA-4), programmed cell death-1 (PD-1), and programmed death-ligand 1 (PD-L1) are widely used as ICIs, and the incidence of irAEs varies depending on the class of agents (3).

Endocrine organs are commonly involved with irAEs, including thyroid irAEs, pituitary irAEs, and diabetes mellitus. Thyroid irAEs are frequently caused by antibodies against PD-1 and PD-L1, but are rarely caused by anti-CTLA-4 antibodies alone (4). Pituitary irAEs are more commonly associated with anti-CTLA-4 antibodies, but can also occur with antibodies against PD-1 and PD-L1 (4). Although diabetes mellitus is a rare irAE associated with antibodies against PD-1 and PD-L1, it requires attention due to its rapid progression to an insulin-dependent condition, which can be life-threatening if not diagnosed.

Combination immunotherapy targeting both the PD-1 and CTLA-4 pathways has been approved and widely adopted for the treatment of cancers such as malignant melanoma, renal cell carcinoma, and non-small cell lung cancer. This combination immunotherapy increases the risk of pituitary irAEs compared to monotherapy targeting only the PD-1 pathway. The incidence of pituitary irAEs due to anti-PD-1 or anti-PD-L1 monotherapy has been reported to be approximately 0.4% in clinical trials (5) and 0.5– 5.7% in cohort studies (6-11). The most common type of pituitary hormone deficiency associated with pituitary irAEs due to PD-1 blockade therapy is isolated ACTH deficiency (IAD) (8,12,13). In contrast, several studies have reported a high incidence of pituitary irAEs with combination immunotherapy using nivolumab, an anti-PD-1 antibody, and ipilimumab, an anti-CTLA-4 antibody, ranging from 6.0% to 29.2% (7,9,10). Moreover, both IAD and combined pituitary hormone deficiency (CPHD) have been observed with this combination immunotherapy (7,9,10). However, detailed information on pituitary irAEs, particularly the differences between CPHD and IAD, remains unclear.

In the present study, we aimed to clarify the clinical features of pituitary irAEs induced by combination immunotherapy with nivolumab and ipilimumab (Nivo/Ipi). Our cohort of patients receiving this therapy revealed that pituitary irAEs include both CPHD and IAD, with the characteristics of CPHD being distinct from those of IAD.

## Materials and Methods

### Patients

This retrospective cohort study was performed using the medical records of consecutive patients who started nivolumab monotherapy (Nivo group) and combination immunotherapy with nivolumab and ipilimumab (Nivo/Ipi group) at Kyoto University Hospital between July 1, 2014 and October 31, 2022. In the Nivo group, patients who received nivolumab as the initial immunotherapy were only included, similarly to our previous studies (8,11,14,15). We excluded patients treated with tyrosine kinase inhibitors or chemotherapy in combination with ICIs, as these agents may affect the occurrence of pituitary irAEs, which has been reported for thyroid irAEs (11,16-18). Since the number of cases was quite small, patients with Hodgkin’s lymphoma, urothelial carcinoma, or cancer of unknown primary origin in the Nivo group, and a patient with colorectal cancer in the Nivo/Ipi group, were excluded. Some patients in the Nivo group were also included in our previous reports (8,14,15).

All their data were collected and fully anonymized in a secured electronic medical record system, and then the dataset was imported to a computer for analysis. This study was approved by the Institutional Review Board and Ethics Committee of the Kyoto University Graduate School of Medicine (approval number, R4981), and was conducted in accordance with the principles of the Declaration of Helsinki. Instead of obtaining informed consent, we provided each patient with the opportunity to opt out of the study using our website.

### Data collection and assays

We collected data from medical records, including age, sex, history of ICI use, cancer type, pituitary magnetic resonance imaging (MRI) results, history of all irAEs, and blood test results. Blood test results included eosinophil count, serum sodium, casual plasma glucose, ACTH, cortisol, TSH, free T4 (fT4), free T3 (fT3), GH, IGF-1, LH, FSH, prolactin (PRL), estradiol, and testosterone. All patient information was followed for at least 6 months.

Plasma ACTH levels were measured using the Elecsys kit (Catalog number 518308322; Roche Diagnostics, Mannheim, Germany), and serum cortisol levels were using the AIA kit (Catalog number 0029138; Tosoh, Tokyo, Japan). Thyroid function tests, including serum levels of TSH, fT4, and fT3, were performed using the Elecsys kits (Catalog number 518315856, 518313180, and 518302702, respectively; Roche Diagnostics): the AIA kits (Catalog number 0029101, 0029103, and 0029102, respectively; Tosoh) were used for a period limited to January 1, 2022, to October 31, 2022. Serum levels of IGF-1 were measured using the Elecsys kit (Catalog number 518313418; Roche Diagnostics) and those of GH, LH, FSH, PRL, estradiol, and testosterone were measured using the the AIA kits (Catalog number 0029135, 0029124, 0029127, 0029177, and 0029130, respectively; Tosoh).

### Assessments of irAEs

Pituitary irAEs were diagnosed by physicians based on clinical symptoms and the confirmation of pituitary hormone deficiency, with reference to the 2023 Guidelines for hypothalamic pituitary dysfunction issued by the Japan Endocrine Society (19). Briefly, ACTH deficiency was diagnosed based on low early morning cortisol and ACTH levels, or a reduced ACTH and cortisol response to the CRH stimulation test. TSH deficiency was identified by low fT4 levels without a compensatory increase in TSH, or a reduced TSH response to the TRH stimulation test. LH/FSH deficiency was diagnosed based on low estradiol or testosterone levels without a compensatory increase in LH and FSH, or a reduced LH and FSH response to the LHRH stimulation test. PRL deficiency was determined by low basal PRL levels, or a reduced PRL response to the TRH stimulation test. GH deficiency was not evaluated using a stimulation test, as no cases showed decreased IGF-1 levels compared to the Japanese reference values (20). Pituitary swelling on MRI was defined as a pituitary height exceeding 10 mm or at least twice the baseline measurement.

Thyroid irAEs were diagnosed according to our previously reported criteria, with a focus exclusively on overt thyroid irAEs, namely situations where neither serum TSH nor fT4 levels were normal (14). Insulin-dependent diabetes as an irAE was defined as hyperglycemia accompanied by low serum C-peptide levels, exclusion of other potential causes, and a sustained requirement for insulin therapy. Non-endocrine irAEs were defined as adverse events affecting organs other than the endocrine system that required intravenous or oral glucocorticoid therapy (equivalent to ≥10 mg of prednisolone) for resolution, in line with our previous study (14).

### Statistical analysis

The number of treated patients during the study period determined the sample size. Data of continuous variables were expressed as medians (Interquartile range: IQR). The Pearson’s chi-square test was used to compare categorical data. The Mann-Whitney U test or the Steel-Dwass test was used for continuous variables. P-values of ≤ 0.05 were considered to indicate statistical significance. JMP Pro 18.0 (SAS Institute Inc., NC, USA) was used to perform the statistical analyses.

## Results

### Patient characteristics and incidence of irAEs

The study included 550 patients in the Nivo group and 92 patients in the Nivo/Ipi group. Patient characteristics and the incidence of irAEs are presented in Table 1. The median age and ratio of male patients were similar between the groups: 69 (62–75) years and 66.5% in the Nivo group, and 69 (58–74) years and 70.7% in the Nivo/Ipi group. Lung cancer and malignant melanoma were the most common primary sites in both groups: 40.5% and 17.3% in the Nivo group, and 35.9% and 37.0% in the Nivo/Ipi group, respectively.

**Table 1.** Patient characteristics in the Nivo and Nivo/Ipi groups.

|  | **Nivo (n = 550)** | **Nivo/Ipi (n = 92)** | **p** |
| --- | --- | --- | --- |
| Age (year) | 69 (62–75) | 69 (58–74) | 0.280 |
| Gender (n) |  |  | 0.438 |
| Male | 366 (66.5%) | 65 (70.7%) |  |
| Female | 184 (33.5%) | 27 (29.3%) |  |
| Primary sites (n) |  |  | **<0.001** |
| Malignant melanoma | **95 (17.3%)** | **34 (37.0%)** |  |
| Lung cancer | **223 (40.5%)** | **33 (35.9%)** |  |
| Others | **232 (42.2%)** | **25 (27.1%)** |  |
| Renal cell carcinoma | 44 (8.0%) | 13 (14.1%) |  |
| Esophageal cancer | 51 (9.3%) | 7 (7.6%) |  |
| Malignant pleural mesothelioma | 7 (1.3%) | 5 (5.4%) |  |
| Gastric cancer | 77 (14.0%) | 0 (0.0%) |  |
| Head and neck carcinoma | 53 (9.6%) | 0 (0.0%) |  |
| irAE (n) |  |  |  |
| Pituitary irAE | **15 (2.7%)** | **16 (17.4%)** | **<0.001** |
| Thyroid irAE | 63 (11.5%) | 13 (14.1%) | 0.462 |
| Thyrotoxicosis and hypothyroidism | 21 (3.8%) | 3 (3.3%) |  |
| Thyrotoxicosis only | 13 (2.4%) | 5 (5.4%) |  |
| Hypothyroidism only | 28 (5.1%) | 5 (5.4%) |  |
| Diabetes mellitus | 5 (0.9%) | 3 (3.3%) | 0.060 |
| Non-endocrine irAE | **65 (11.8%)** | **33(35.9%)** | **<0.001** |
| Pneumonitis | 31 (5.6%) | 6 (6.5%) |  |
| Dermatopathy | 7 (1.3%) | 4 (4.3%) |  |
| Colitis | 6 (1.1%) | 5 (5.4%) |  |
| Hepatitis | 4 (0.7%) | 10 (10.9%) |  |
| Acute kidney injury | 4 (0.7%) | 2 (2.2%) |  |
| Others | 13 (2.4%) | 12 (13.0%) |  |

Nivo, nivolumab monotherapy; Nivo/Ipi, combination therapy using nivolumab and ipilimumab; n, number of subjects; irAE, immune-related adverse event. ‘Others’ in non-endocrine irAE includes the following events: Polymyalgia rheumatica (n = 3), myocarditis (n = 2), ITP (n = 1), scleroderma (n = 1), mucosal rash (n = 1), arthritis (n = 1), uveitis (n = 1), gastritis (n = 1), headache (n = 1), and myositis (n = 1) in the Nivo group: and fever (n = 3), esophagitis (n = 2), myositis (n = 2), polymyalgia rheumatica (n = 1), peripheral neuropathy (n = 1), myocarditis (n = 1), laryngitis (n = 1), and
pancreatitis (n = 1) in the Nivo/Ipi group. Data of continuous variables are expressed as medians (interquartile range). Statistical analyses were performed using the Mann-Whitney U test for continuous variables and the Pearson's chi-square test for categorical variables. Significant differences are shown in bold.

The incidence of pituitary irAEs was significantly higher in the Nivo/Ipi group (16 of 92 patients, 17.4%) than in the Nivo group (15 of 550 patients, 2.7%) (p < 0.001). On the other hand, thyroid irAEs and diabetes mellitus occurred at similar rates in the groups: 11.5% and 0.9% in the Nivo group, and 14.1% and 3.3% in the Nivo/Ipi group, respectively. Similar to pituitary irAEs, non-endocrine irAEs were significantly more frequent in the Nivo/Ipi group (33 of 92 patients, 35.9%) than in the Nivo group (65 of 550 patients, 11.8%) (p < 0.001). Among non-endocrine irAEs, dermatopathy, colitis, and hepatitis occurred more frequently in the Nivo/Ipi group (4.3%, 5.4%, and 10.9%, respectively) compared to the Nivo group (1.3%, 1.1%, and 0.7%, respectively), which is consistent with previous reports (21,22).

According to the inclusion criteria, no patients in the Nivo group had received prior ICIs. In contrast, 31 of 92 patients (34.8%) in the Nivo/Ipi group had previously received ICIs: 15 with nivolumab, 10 with pembrolizumab, 3 with durvalumab, 1 with atezolizumab, 1 with both nivolumab and pembrolizumab, and 1 with both pembrolizumab and atezolizumab. However, no significant differences in the incidence of irAEs were observed based on prior ICI use (Supplementary Table 1).

### Clinical features of pituitary irAEs with and without ipilimumab use

As mentioned above, the incidence of pituitary irAEs was higher in the Nivo/Ipi group than in the Nivo group. To elucidate differences in clinical features, we compared data on pituitary irAEs between the groups. Pituitary irAEs developed significantly earlier in the Nivo/Ipi group (Median, 63 days; IQR, 29–89) compared to the Nivo group (Median, 154 days; IQR, 115–273) (p < 0.001) (Figure 1A). Detailed data of all patients who developed pituitary irAEs are presented in Supplementary Tables 2 and 3. With regard to patterns of pituitary dysfunction, all 15 patients in the Nivo group exhibited IAD, whereas in the Nivo/Ipi group, 8 patients (50.0%) had IAD and 8 patients (50.0%) had CPHD (p < 0.001) (Figure 1B). Among the patients with CPHD in the Nivo/Ipi group, ACTH deficiency was observed in all 8 patients (100.0%), LH/FSH deficiency in 6 of 8 (75.0%), TSH deficiency in 5 of 8 (62.5%), and PRL deficiency in 3 of 8 (37.5%). GH deficiency was not observed, as IGF-1 levels remained within the reference range. ACTH deficiency did not restore in any patient, but partial restoration was observed for other hormones: LH/FSH in 3 of 5 (60.0%), TSH in 3 of 4 (75.0%), and PRL in 2 of 2 (100.0%) reevaluated patients (Supplementary Table 3).

**Figure 1.**
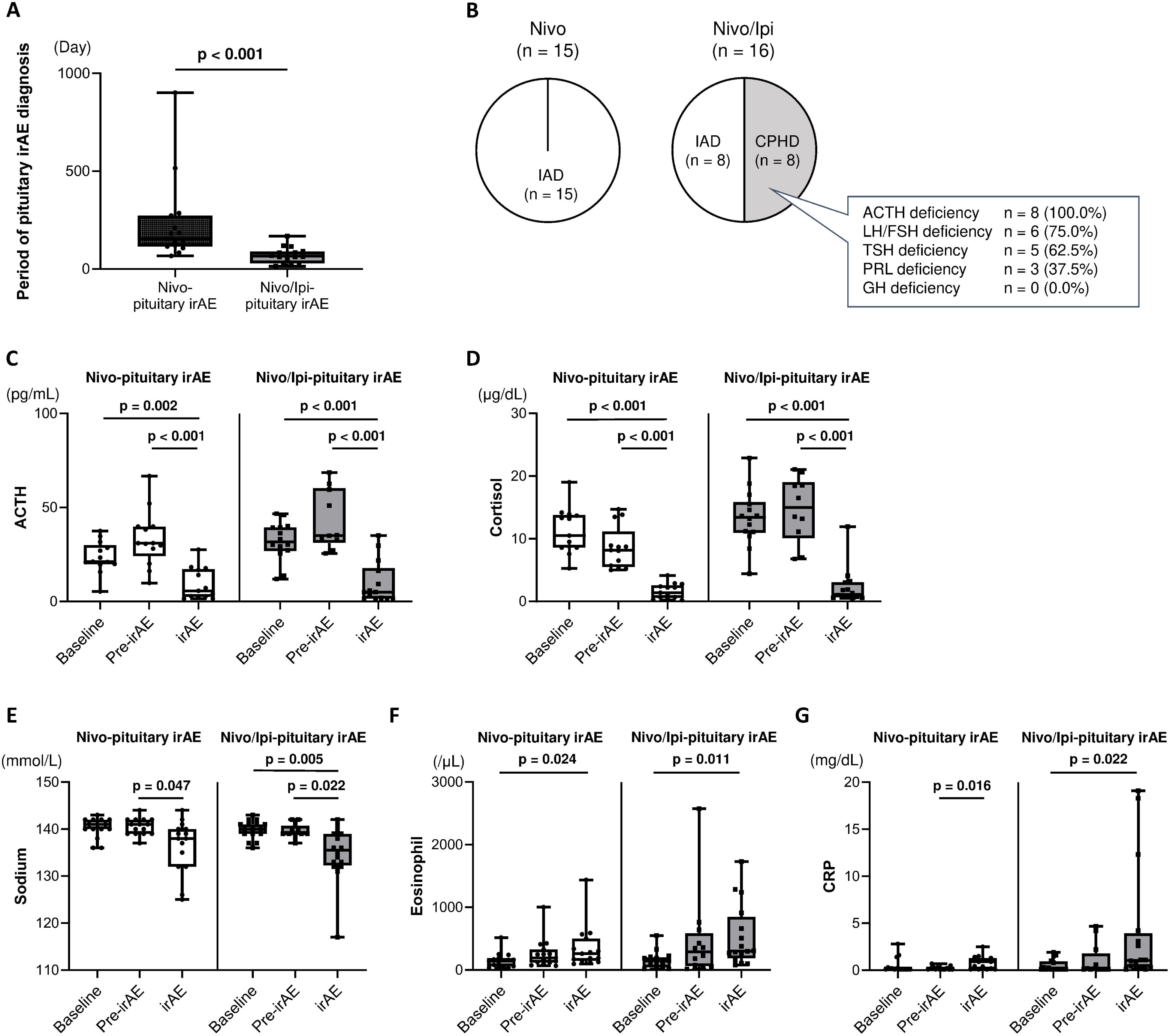
Clinical features of pituitary irAEs with and without ipilimumab treatment. (A) Period of pituitary irAE diagnosis. ‘Day’ is expressed as the number of days since the first administration of nivolumab (Nivo) or nivolumab and ipilimumab (Nivo/Ipi). irAE, immune related adverse event. (B) Patterns of pituitary dysfunction. IAD, isolated ACTH deficiency; CPHD, combined pituitary hormone deficiency; PRL, prolactin. (C–G) Longitudinal changes in clinical parameters at three time points, baseline, the last measurement before pituitary irAE (pre-irAE), and at pituitary irAE diagnosis (irAE): plasma ACTH levels (C), serum cortisol levels (D), serum sodium levels (E), eosinophil counts (F), and serum C-reactive protein (CRP) levels (G). Statistical analyses were performed using the Mann-Whitney U test for panel A, and the Steel-Dwass test for panels C–G. Significant differences (p < 0.05) are highlighted in boldface text, and results for p ≥ 0.05 are not displayed.

In these patients with pituitary irAEs, the incidence of thyroid irAEs was similar between the two groups (Supplementary Figure 1A). On the other hand, non-endocrine irAEs were more frequently observed in the Nivo/Ipi group (6 of 16 patients, 37.5%) than in the Nivo group (1 of 14 patients, 6.7%) (p = 0.040) (Supplementary Figure 1B).

We analyzed the longitudinal laboratory data of patients who developed pituitary irAEs. In both groups, circulating levels of ACTH and cortisol showed a similar trend. Plasma ACTH levels remained unchanged at a period just before the diagnosis of pituitary irAEs (pre-irAE) but significantly decreased at irAE diagnosis compared to baseline (Figure 1C): in the Nivo group, ACTH levels were 21.2 (19.9– 30.2) pg/mL at baseline, 31.0 (24.1–39.9) pg/mL at pre-irAE, and 5.7 (2.5–17.3) pg/mL at irAE diagnosis (p = 0.002 for baseline vs. irAE diagnosis); and in the Nivo/Ipi group, 31.7 (26.8–39.5) pg/mL at baseline, 35.1 (31.2–60.3) pg/mL at pre-irAE, and 5.0 (1.5– 17.8) pg/mL at irAE diagnosis (p < 0.001 for baseline vs. irAE diagnosis). Serum cortisol levels followed the same pattern as ACTH (Figure 1D): in the Nivo group, cortisol levels were 10.5 (8.6–13.8) μg/dL at baseline, 8.2 (5.5–11.2) μg/dL at pre-irAE, and 1.4 (0.6–2.6) μg/dL at irAE diagnosis (p < 0.001 for baseline vs. irAE diagnosis); and in the Nivo/Ipi group, 13.4 (10.9–15.9) μg/dL at baseline, 15.0 (10.1–19.0) μg/dL at pre-irAE, and 1.2 (0.7–3.1) μg/dL at irAE diagnosis (p < 0.001 for baseline vs. irAE diagnosis).

Serum sodium levels showed similar trends in both groups, remaining stable at pre-irAE and decreasing at irAE diagnosis (Figure 1E). Eosinophil counts gradually increased from baseline to irAE diagnosis in both groups (Figure 1F). Serum C-reactive protein (CRP) levels exhibited different trends between the Nivo and Nivo/Ipi groups (Figure 1G). In the Nivo group, there was no significant change between baseline and irAE diagnosis. In contrast, in the Nivo/Ipi group, CRP levels remained unchanged at pre-irAE but increased at irAE diagnosis: 0.1 (0.1–1.0) mg/dL at baseline, 0.1 (0.1–1.8) mg/dL at pre-irAE, and 1.1 (0.4–4.0) mg/dL at irAE diagnosis (p = 0.022 for baseline vs. irAE diagnosis).

We previously reported that IAD induced by nivolumab monotherapy was accompanied by thyroid dysfunction, characterized by an elevated fT3/fT4 ratio and increased fT3 levels without TSH suppression (8). Based on this finding, we further investigated thyroid function in patients with pituitary irAEs. In the Nivo group, serum fT3 levels increased at irAE onset: 2.68 (2.35–3.04) pg/mL at baseline, 2.73 (2.33–3.09) pg/mL at pre-irAE, and 3.48 (3.06–3.68) pg/mL at irAE diagnosis (p = 0.020 for pre-irAE vs. irAE diagnosis) (Supplementary Figure 2A). Serum fT4 and TSH levels remained unchanged, whereas fT3/fT4 ratio significantly increased at irAE diagnosis: 2.36 (2.26–2.77) at baseline, 2.71 (2.55–2.94) at pre-irAE, and 3.55 (3.09–4.10) at irAE diagnosis (p = 0.031 for baseline vs. irAE; p = 0.045 for pre-irAE vs. irAE diagnosis) (Supplementary Figure 2B–D). In the Nivo/Ipi group, serum fT3 and TSH levels remained unchanged, while serum fT4 levels decreased at irAE diagnosis: 1.15 (1.05– 1.20) ng/dL at baseline, 1.10 (1.04–1.26) ng/dL at pre-irAE, and 0.69 (0.54–1.06) ng/dL at irAE diagnosis (p = 0.047 for baseline vs. irAE diagnosis) (Supplementary Figure 2A–C). In addition, fT3/fT4 ratio significantly increased at irAE diagnosis: 2.18 (1.64– 2.44) at baseline, 2.03 (1.54–2.44) at pre-irAE, and 3.34 (2.32–4.19) at irAE diagnosis (p = 0.048 for baseline vs. irAE diagnosis) (Supplementary Figure 2D). Thus, the pattern of thyroid function changes associated with pituitary irAEs differed between the groups.

### Comparison between CPHD and IAD induced by Nivo/Ipi therapy

To clarify the characteristics of pituitary irAEs induced by combination immunotherapy, we analyzed data from the Nivo/Ipi group by stratifying patients into those with CPHD (n = 8) and those with IAD (n = 8). Pituitary irAEs developed significantly earlier in patients with CPHD (Median, 40 days; IQR, 20–73) compared to those with IAD (Median, 84 days; IQR., 62–121) (p = 0.022) (Figure 2A). Patients with CPHD received significantly fewer ICI administrations (Median, 2; IQR, 1–3) compared to those with IAD (Median, 3; IQR, 1–4) (p = 0.032) (Figure 2B). Notably, 4 of 8 patients with CPHD were diagnosed with pituitary irAEs during the initial course of Nivo/Ipi administration (Cases 9, 10, 12, and 16 in Supplementary Table 3). There was no significant difference in the number of patients who had received prior ICI treatment between the two groups (4 of 8 patients with CPHD vs. 2 of 8 patients with IAD, p = 0.302).

**Figure 2.**
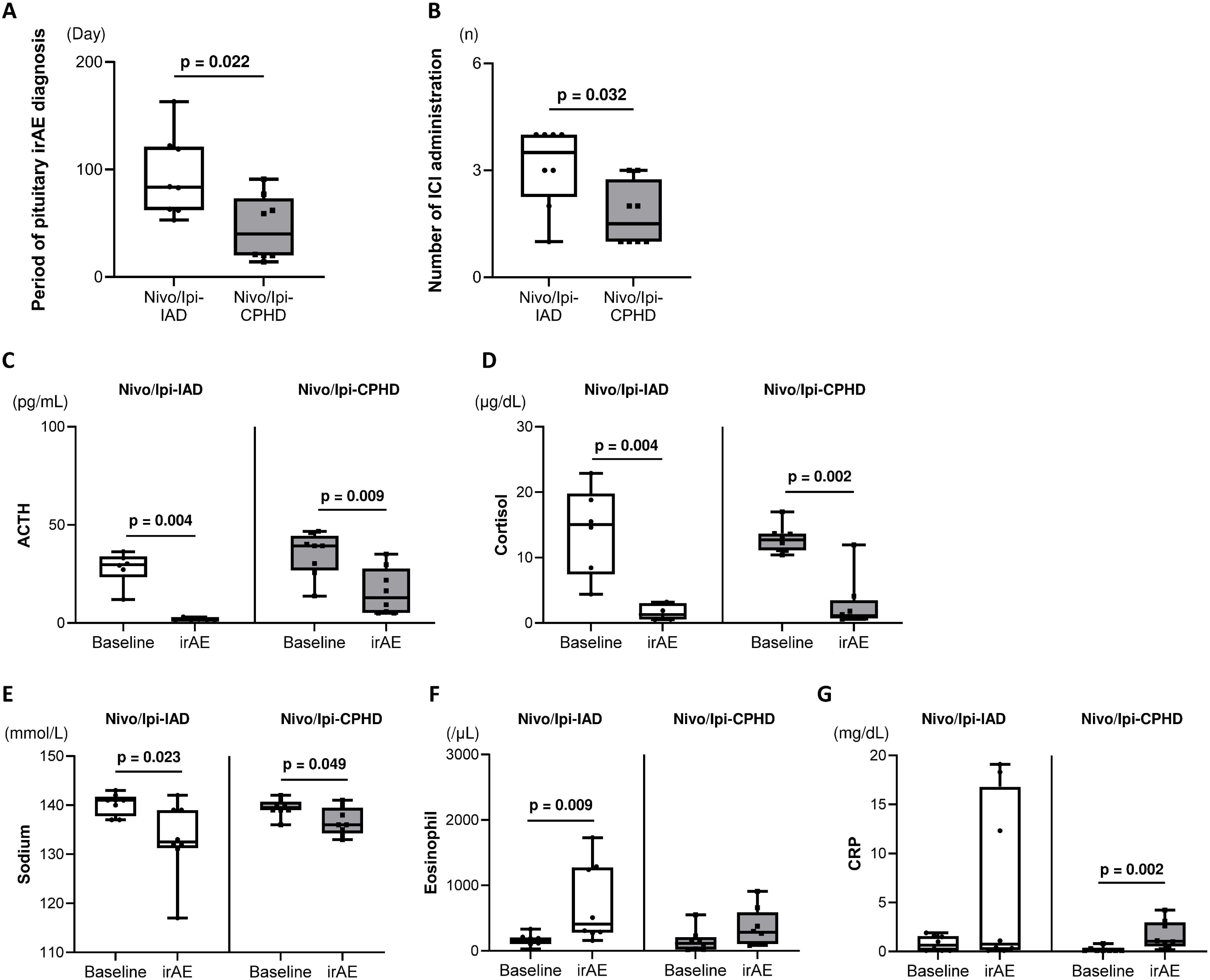
Comparisons of clinical features between CPHD and IAD induced by Nivo/Ipi therapy. (A) Period of pituitary irAE diagnosis. (B) Number of ICI administrations before pituitary irAE diagnosis. (C–G) Longitudinal changes in clinical parameters at baseline and at pituitary irAE diagnosis (irAE): plasma ACTH levels (C), serum cortisol levels (D), serum sodium levels (E), eosinophil counts (F), and serum CRP levels (G). Statistical analyses were performed using the Mann–Whitney U test. Significant differences are displayed only.

We evaluated the longitudinal changes in laboratory data, focusing on CPHD and IAD. Only data at baseline and irAE diagnosis were analyzed, because half of patients with CPHD developed pituitary irAEs during the initial course. Plasma ACTH levels significantly decreased at irAE diagnosis compared to baseline (Figure 2C): in patients with IAD, 29.8 (23.4–33.9) pg/mL at baseline to 1.5 (1.5–2.2) pg/mL at irAE diagnosis (p = 0.004);and in patients with CPHD, ACTH levels dropped from 39.3 (26.8–44.5) pg/mL at baseline and 12.9 (5.2–27.8) pg/mL at irAE diagnosis (p = 0.009). Serum cortisol levels showed a similar trend (Figure 2D): in patients with IAD, 15.1 (7.4–19.8) μg/dL at baseline and 1.3 (0.6–3.1) μg/dL at irAE diagnosis (p = 0.004); and in patients with CPHD, 12.7 (11.1–13.7) μg/dL at baseline and 1.2 (0.7–3.5) μg/dL at irAE diagnosis (p = 0.002).

Serum sodium levels significantly decreased at irAE diagnosis in both groups, similar to ACTH and cortisol levels (Figure 2E): in patients with IAD, 141 (138–142) mEq/L at baseline and 133 (131–139) mEq/L at irAE diagnosis (p = 0.023); in patients with CPHD, 140 (139–141) mEq/L at baseline and 136 (134–140) mEq/L at irAE diagnosis (p = 0.049). In contrast, eosinophil counts and CRP levels exhibited distinct patterns between the groups. Eosinophil counts significantly increased in patients with IAD, from 166 (105–206) /μL at baseline to 409 (276–1275) /μL at irAE diagnosis (p = 0.009), whereas in patients with CPHD, levels remained unchanged (Figure 2F). Similarly, CRP levels remained largely unchanged in patients with IAD (Figure 2G), although marked elevations were observed in some patients (Supplementary Table 3; cases 3, 5, and 6). In contrast, CRP levels significantly increased in a large proportion of patients with CPHD, rising from 0.1 (0.1–1.0) mg/dL at baseline to 1.1 (0.1–4.0) mg/dL at irAE diagnosis (p = 0.002) (Figure 2G).

Thyroid function also exhibited distinct trends between the groups. In patients with IAD, fT3, fT4, and TSH levels remained unchanged, whereas fT3/fT4 ratio significantly increased at irAE diagnosis compared to baseline (Supplementary Figure 3A–D): from 2.05 (1.64–2.52) at baseline to 3.20 (2.05–4.13) at irAE diagnosis (p = 0.010) (Supplementary Figure 3D). In patients with CPHD, fT4 and TSH levels significantly decreased: fT4 declined from 1.09 (1.01–1.30) ng/dL at baseline to 0.54 (0.50–0.60) ng/dL at irAE diagnosis (p = 0.030) (Supplementary Figure 3B), and TSH decreased from 4.450 (2.690–6.341) μIU/mL at baseline to 0.482 (0.225–1.906) μIU/mL at irAE diagnosis (p = 0.030) (Supplementary Figure 3C).

Adrenal insufficiency-related symptoms, such as fatigue and loss of appetite, did not significantly differ between patients with CPHD and those with IAD (Figure 3A). Fever was observed only in patients with CPHD (2 of 8 patients, 25.0%), and headaches were significantly more frequent in patients with CPHD (6 of 8 patients, 75.0%) than in those with IAD (1 of 8 patients, 12.5%) (p = 0.012) (Figure 3A). We also examined pituitary MRI results (Supplementary Figure 4), with representative images shown in Figure 3B. Pituitary swelling was observed in a large proportion of patients with CPHD (6 of 8 patients, 75.0%) whereas none of the patients with IAD exhibited this finding (0 of 5 patients, 0.0%) (p = 0.008) (Figure 3C). The incidence of thyroid irAEs was similar between the groups; however, the incidence of non-endocrine irAEs was significantly higher in patients with CPHD (6 of 8 patients, 75.0%) than in those with IAD (0 of 8 patients, 0.0%) (p = 0.002) (Figure 3D).

**Figure 3.**
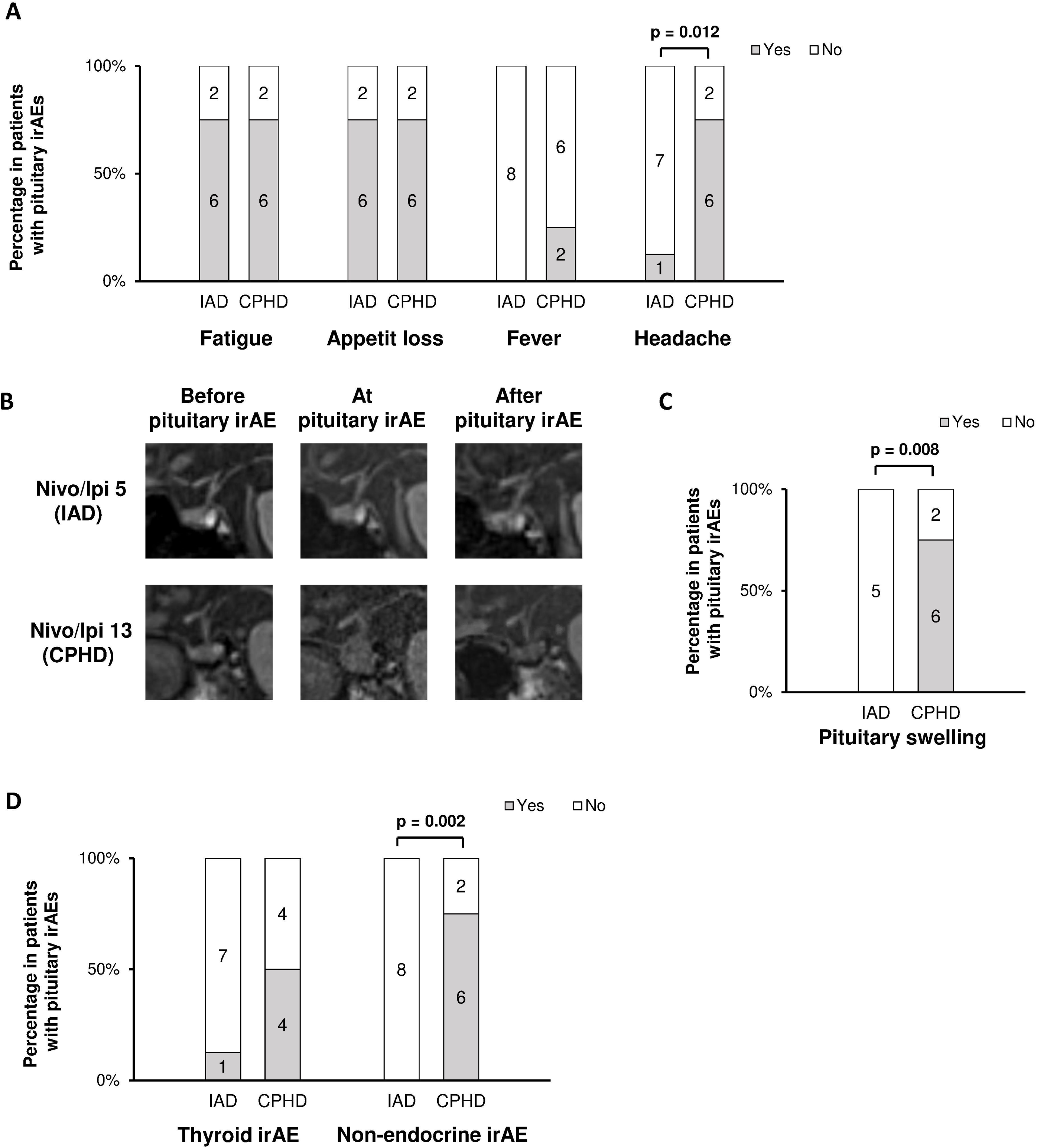
Comparisons of symptoms, MRI findings, and concomitant irAEs between CPHD and IAD induced by Nivo/Ipi therapy. (A) Incidence of symptoms associated with pituitary irAEs. (B) Representative magnetic resonance imaging (MRI) findings in cases of IAD and CPHD at three time points: before the onset of pituitary irAE (before pituitary irAE), at the time of pituitary irAE diagnosis (at pituitary irAE), and after the onset of pituitary irAE (after pituitary irAE). (C) Frequency of pituitary swelling. (D) Incidence of thyroid irAEs and non-endocrine irAEs. Their detailed clinical information is provided in Supplementary Table 3. Statistical analyses were performed using the Chi-square test. Significant differences are displayed only.

A substantial number of patients with pituitary irAEs in the Nivo/Ipi group had malignant melanoma (5 of 8 patients with CPHD and 4 of 8 patients with IAD). A previous report suggests that pituitary irAEs occur more frequently in malignant melanoma than in renal cell carcinoma (9). In Japan, ipilimumab is administered at a higher dose (3 mg/kg) for malignant melanoma compared to the lower dose (1 mg/kg) used for other cancers. Therefore, we further analyzed the Nivo/Ipi group based on the administered dose of ipilimumab. The 3 mg/kg group presented a trend toward a higher incidence of pituitary irAEs (9 of 34 patients, 26.5%) compared to the 1 mg/kg group (7 of 58 patients, 12.1%) (p = 0.079) (Supplementary Table 4). There were no significant differences in the incidence of thyroid irAEs between the groups (Supplementary Figure 5A). Non-endocrine irAEs were significantly more frequent in the 3 mg/kg group (21 of 34 patients, 61.8%) than in the 1 mg/kg group (12 of 58 patients, 20.7%) (p < 0.001) (Supplementary Figure 5B). Specifically, dermatopathy, colitis, and hepatitis were more frequent in the 3 mg/kg group (11.8%, 14.7%, and 26.5%, respectively) compared to the 1 mg/kg group (0.0%, 0.0%, and 1.7%, respectively) (Supplementary Table 4).

### Lessons from recurrent cases of CPHD

As described above, CPHD and IAD observed during Nivo/Ipi therapy exhibited distinct clinical characteristics. Pituitary irAEs are generally treated with physiological doses of hydrocortisone, except in cases with chiasmal compression or severe headache (23,24). Therefore, in IAD cases without pituitary swelling, physiological doses of glucocorticoids are generally sufficient. On the other hand, to highlight the clinical course and management challenges of CPHD, we present two cases in which pituitary irAEs worsened following Nivo/Ipi re-administration.

The first case (Figure 4A, Case 9 in Supplementary Table 3) underwent surgical resection for malignant melanoma and subsequently received adjuvant pembrolizumab monotherapy 13 times at 3-week intervals. Metastases to the lungs, liver, and gastric mucosa prompted the initiation of combination immunotherapy. On day 14 after the first Nivo/Ipi administration, he reported fatigue, appetite loss, and headache. Laboratory tests confirmed CPHD, including ACTH and PRL deficiencies (Table 2). Hydrocortisone was initiated at a dose of 20 mg/day, which led to symptomatic improvement. However, MRI revealed persistent pituitary swelling despite hydrocortisone treatment (Figure 4A). The second Nivo/Ipi administration was performed on day 28, after which he experienced worsening headaches and newly developed LH/FSH deficiency (Table 2). The hydrocortisone dose was increased to 40 mg/day, resulting in partial relief of headache. On day 49, he developed additional irAEs, including pharyngitis and gastric mucosal disorder, for which prednisolone (PSL) was initiated. The headache completely resolved, and follow-up MRI showed improvement in pituitary swelling (Figure 4A). LH/FSH and PRL deficiencies also resolved (Table 2).

**Figure 4.**
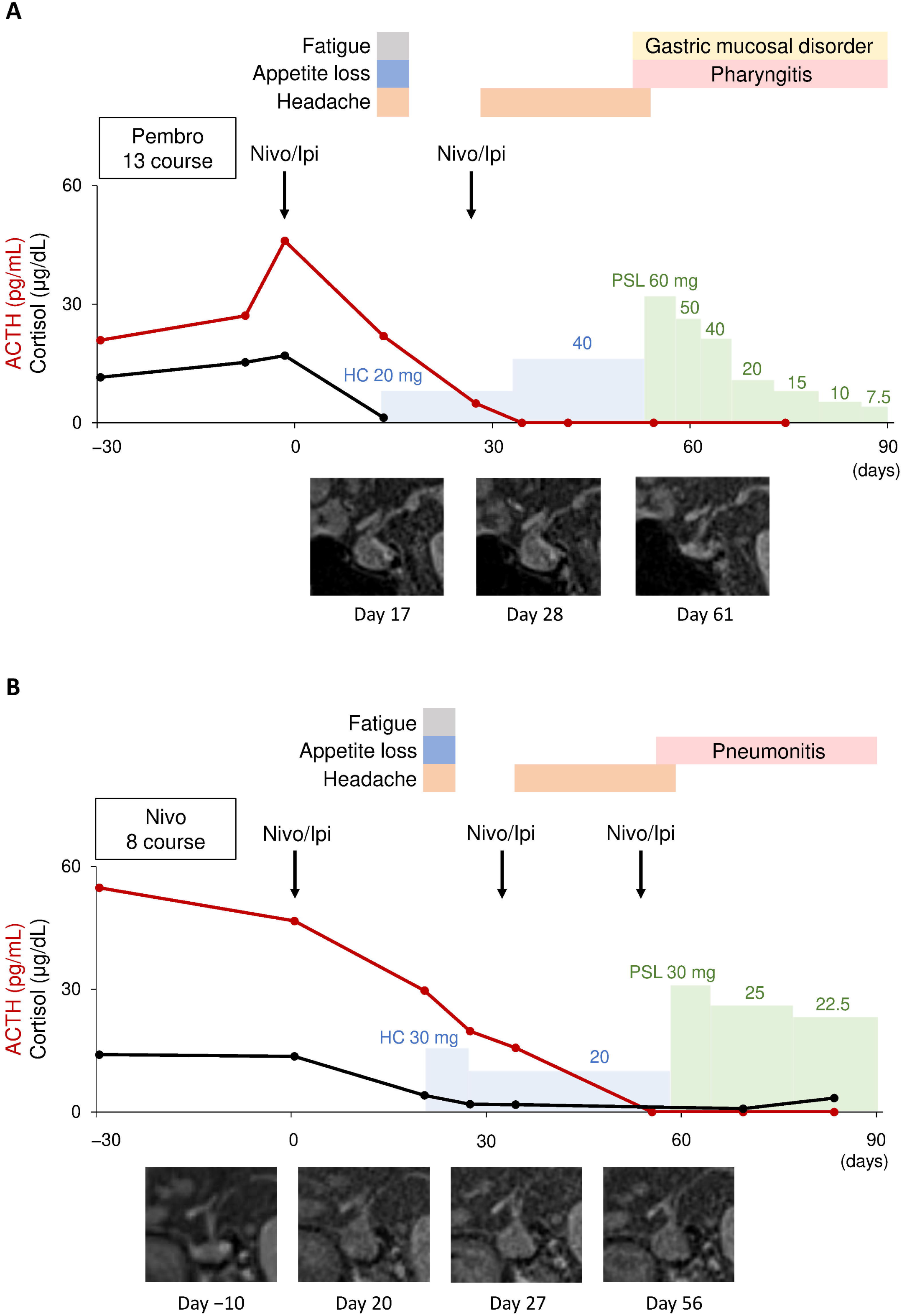
Clinical courses of recurrent CPHD following Nivo/Ipi re-administration. (A) Detailed information is provided as Case 9 in Supplementary Table 3. (B) Detailed information is provided as Case 13 in Supplementary Table 3. Day 0 represents the date of the first Nivo/Ipi administration. Pembro, pembrolizumab; HC, hydrocortisone; PSL, prednisolone.

**Table 2.** Longitudinal data on cases with CPHD presented in Figure 4.

| Day | Case 1 (Figure 4A) |  |  |  |  |  |  | Case 2 (Figure 4B) |  |  |  |  |  |  |
| --- | --- | --- | --- | --- | --- | --- | --- | --- | --- | --- | --- | --- | --- | --- |
|  | -2 | 14 | 28 | 35 | 42 | 55 | 75 | -6 | 0 | 20 | 34 | 55 | 69 | 83 |
| ftT3 (pg/mL) | 2.43 | 3.71 | 2.78 | 2.15 | 2.25 | 2.53 | 2.55 | 1.98 | 2.05 | 1.61 | 1.58 | 1.20 | 1.30 | 1.46 |
| ftT4 (ng/dL) | 1.15 | 1.33 | 1.15 | 1.14 | 1.39 | 3.46 | 1.60 | 0.80 | 1.02 | 0.53 | 0.79 | 0.69 | 1.32 | 1.40 |
| TSH (μIU/mL) | 0.296 | 0.092 | <0.005 | <0.005 | <0.005 | <0.005 | <0.005 | 13.613 | 6.585 | 0.150 | 0.192 | 0.016 | 0.117 | 0.156 |
| GH (ng/mL) | NA | 1.47 | 1.13 | 0.86 | 1.89 | 0.92 | NA | NA | NA | 1.54 | 0.41 | 0.12 | 0.10 | 0.11 |
| IGF-1 (ng/mL) | NA | 210 | 258 | 216 | 186 | 170 | NA | NA | NA | 87 | 118 | 49 | 48 | 62 |
| PRL (ng/mL) | NA | 3.5 | 3.0 | 2.8 | 3.6 | 5.6 | 8.6 | NA | NA | 7.4 | 9.3 | 1.4 | 5.8 | 7.1 |
| LH (mIU/mL) | NA | 5.8 | 0.8 | 2.0 | 2.9 | 7.5 | 6.1 | NA | NA | 1.5 | 3.7 | 0.9 | 3.4 | 5.7 |
| FSH (mIU/mL) | NA | 22.2 | 2.3 | 3.8 | 4.2 | 8.0 | 9.0 | NA | NA | 7.0 | 8.8 | 3.7 | 9.6 | 14.3 |
| T (ng/dL) | NA | 216.3 | <2.5 | 47.3 | 150.1 | 595.0 | NA | NA | NA | <7.0 | 119.4 | <7.0 | <7.0 | 32.7 |
| HC (mg/day) | 0 | 20 | 40 | 40 | 40 | 0 | 0 | 0 | 0 | 30 | 20 | 20 | 0 | 0 |
| PSL (mg/day) | 0 | 0 | 0 | 0 | 0 | 60 | 15 | 0 | 0 | 0 | 0 | 0 | 30 | 22.5 |
| LT4 (μg/day) | 0 | 0 | 0 | 0 | 0 | 0 | 0 | 25 | 25 | 25 | 25 | 25 | 50 | 50 |

CPHD, combined pituitary hormone deficiency; ftT3, free T3; free T4, ftT4; PRL, prolactin; T, testosterone; HC, hydrocortisone; PSL, prednisolone; LT4, levothyroxine. 'Days' is expressed as the number of days since the first administration of Nivolumab/Ipilimumab. TSH, ftT4, and ftT3 were measured using the Elecsys kits in Case 1 and the AIA kits in Case 2. Reference ranges: ftT3, 2.33–4.00 pg/mL (Elecsys), 2.06–3.17 pg/mL (AIA); ftT4, 0.880–1.620 ng/dL (Elecsys and AIA); TSH, 0.500–5.000 μIU/mL (Elecsys and AIA); GH, ≤2.10 ng/mL; IGF-1, age- and sex-specific reference ranges (21); PRL, 3.6–16.3 ng/mL; LH, 1.7–8.6 mIU/mL; FSH, 1.5–12.4 mIU/mL; Testosterone (T), 131.0–871.0 ng/dL.

The second case (Figure 4B, Case 13 in Supplementary Table 3) also underwent surgical resection for malignant melanoma and received adjuvant nivolumab monotherapy 8 times at 4-week intervals. Local recurrence, calcaneal metastasis, and inguinal lymph node metastasis prompted the initiation of combination immunotherapy. On day 20 after the first Nivo/Ipi administration, he developed fatigue, appetite loss, and headache, similar to the first case. Laboratory tests confirmed CPHD, including ACTH, TSH, and LH/FSH deficiencies (Table 2). MRI revealed pituitary swelling, which had not been observed before the first Nivo/Ipi administration (Figure 4B). Hydrocortisone was initiated at a dose of 30 mg/day and then reduced to 20 mg/day as symptoms improved. Additionally, TSH and LH/FSH deficiencies showed signs of recovery (Table 2). However, after the second Nivo/Ipi administration on day 32, he experienced worsening headaches along with a recurrence of TSH and LH/FSH deficiencies (Table 2). Following the third Nivo/Ipi administration on day 53, MRI revealed persisted pituitary swelling on day 56 (Figure 4B). On day 57, he developed irAE-related pneumonitis, for which PSL was initiated. The headache rapidly resolved, and LH/FSH and TSH deficiencies gradually improved (Table 2).

To summarize, the above cases of CPHD presented with headache symptoms, which initially improved with hydrocortisone at a dose of 20 mg/day. However, pituitary dysfunction and headaches worsened after the second Nivo/Ipi administration. Following the initiation of pharmacologic doses of PSL for concomitant non-endocrine irAEs, both pituitary dysfunction and headaches improved. These findings suggest that, unlike IAD, physiological doses of hydrocortisone are insufficient to manage certain cases of CPHD during combination immunotherapy.

## Discussion

Through this cohort study, we clarified clinical features of pituitary irAEs induced by combination immunotherapy with Nivo/Ipi. Compared to nivolumab monotherapy, combination immunotherapy resulted in a higher incidence and earlier onset of pituitary irAEs. By analyzing longitudinal data, we identified differences in the trends of laboratory findings, including serum CRP levels and thyroid function. Moreover, we demonstrated that CPHD was observed only with combination immunotherapy and exhibited distinct clinical features compared to IAD. Firstly, CPHD occurred further earlier than IAD. Secondly, headache and pituitary swelling were predominantly observed in CPHD. Thirdly, CPHD was often accompanied by non-endocrine irAEs. The management of CPHD appears challenging, as it can relapse with re-administration of Nivo/Ipi, requiring pharmacological doses of glucocorticoids.

So far, several clinical studies have reported pituitary irAEs induced by combination immunotherapy with PD-1 and CTLA-4 inhibitors. These studies have shown that pituitary irAEs occur more frequently and earlier with combination immunotherapy (6.0–29.2%, 2.0–3.5 months) than with anti-PD-1 or anti-PD-L1 monotherapy (1.1–5.7%, 4.1–5.5 months) (6,7,9,10,25). The validity of our cohort is supported by consistency in the epidemiology: the incidence and onset of pituitary irAEs were 17.4% and 63 days in the Nivo/Ipi group, compared to 2.7% and 154 days in the Nivo group.

The analyses of laboratory data trends identified differences according to ICI regimens. Changes in ACTH, cortisol, sodium, and eosinophil counts were almost similar between the Nivo and Nivo/Ipi groups. However, CRP levels significantly increased in the Nivo/Ipi group, suggesting the occurrence of non-endocrine irAEs. Changes in thyroid function also differed between the groups. In the Nivo group, fT3 and fT3/fT4 increased without a negative feedback response of TSH, consistent with our previous report (8). On the other hand, in the Nivo/Ipi group, the primary change was a decrease in fT4. TSH levels did not increase despite low fT4, suggesting these changes were a consequence of TSH deficiency. Thyroid function may not serve as a reliable reference for pituitary irAEs in combination immunotherapy.

To better understand pituitary irAEs induced by combination immunotherapy, we classified based on patterns of pituitary dysfunction. As previously reported, pituitary irAEs induced by nivolumab monotherapy exclusively exhibited IAD (7,8,25), whereas those induced by combination immunotherapy were associated with both CPHD and IAD (6,7,9,25). In this cohort, CPHD frequently involved LH/FSH deficiency (37.5%, 6 of 16 pituitary irAE cases in the Nivo/Ipi group), and TSH deficiency (31.8%, 5 of 16 pituitary irAE cases in the Nivo/Ipi group), which closely aligns with previous reports (25.0–57.1%, 36.0–85.7%, respectively) (6,7,9,25). PRL deficiency was observed in 3 of 16 pituitary irAE cases (18.8%), which is lower than previously reported rates (57.1–75.0%) (7,10). However, prior studies did not assess PRL levels in all cases, and the reported percentages were based only on those tested, potentially explaining this discrepancy. GH deficiency was not observed in this study, whereas previous reports have noted an incidence of 28.6–33.3% (7,10,25). Since our evaluation was based solely on IGF-1 levels without performing GH stimulation tests, the prevalence of GH deficiency may have been underestimated. Diabetes insipidus was not observed in this study, similar to the previous report (25). Regarding the outcome of pituitary dysfunction, it has been well documented that IAD due to nivolumab monotherapy did not recover in any cases (6,25). In combination immunotherapy, while ACTH deficiency is irreversible, LH/FSH and TSH deficiencies improve in 60–70% and 58–92% of cases, respectively (6,25). These are consistent with our own observation: ACTH deficiency did not recover in any cases of pituitary irAEs, while recovery was noted in 60.0% of LH/FSH deficiency and 75.0% of TSH deficiency.

Even when focusing solely on IAD, it occurred earlier in the Nivo/Ipi group (median 84 days) than that in the Nivo group (median 154 days). The incidence of IAD was higher in the Nivo/Ipi group (8.7%) compared to the Nivo group (2.7%). These findings suggest that CTLA-4 blockade, as well as PD-1 blockade, may contribute to the development of IAD. Notably, CPHD developed further earlier (median 40 days) than IAD (median 84 days) in the Nivo/Ipi group, with 4 patients experiencing onset during the first course. While pituitary irAEs have traditionally been considered to occur a few months after treatment initiation, our findings suggest that in combination immunotherapy, they can develop at an extremely early stage. Therefore, physicians should be highly aware of the risk of pituitary irAEs from the very beginning of treatment.

Headache is highlighted as a key manifestation of CPHD. Fatigue and appetite loss, major symptoms related to adrenal insufficiency, were observed similarly in both CPHD and IAD. Fever was exclusively observed in CHPD, but it occurred less frequently than headaches. From a clinical management perspective, if patients report headaches during combination immunotherapy, physicians should suspect pituitary irAEs. However, this approach cannot be applied when suspecting IAD. Laboratory findings also characterize CPHD. Patients with IAD in both the Nivo and Nivo/Ipi groups showed similar trends in serum sodium levels, eosinophil counts, and thyroid function. The differences observed in CRP levels and thyroid function between pituitary irAEs in the Nivo/Ipi group and those in the Nivo group likely reflect the presence of patients with CPHD.

The appropriate glucocorticoid dosage for CPHD and IAD should be carefully considered. Physiological hydrocortisone replacement is typically sufficient for managing pituitary irAEs, except in cases involving chiasmal compression or severe headache (23,24). In our cohort, regardless of whether pituitary irAEs were induced by Nivo or Nivo/Ipi, none of the IAD cases showed symptoms worsening with physiological doses of hydrocortisone. In contrast, the two CPHD cases experienced clinical worsening following re-administration of Nivo/Ipi, and their symptoms and pituitary dysfunction improved with pharmacological doses of PSL. Therefore, physicians should recognize that some CPHD cases may require higher glucocorticoid doses to prevent the worsening of pituitary irAEs after re-administration of Nivo/Ipi.

Given that CPHD is rarely induced by PD-1 blockade alone, CTLA-4 blockade is likely implicated in its pathogenesis. Anti-CTLA-4 antibodies have been shown to exhibit a dose-dependent relationship with various irAEs, particularly hepatitis and gastrointestinal disorders, which increase in frequency with higher doses of ipilimumab (26). In this study, dermatopathy, colitis, and hepatitis occurred more frequently in patients receiving higher doses of ipilimumab. Furthermore, patients treated with higher doses of ipilimumab tended to be more susceptible to developing pituitary irAEs. Thus, CTLA-4 blockade seems to play a significant role in the development of pituitary irAEs.

From a molecular perspective, CTLA-4 is expressed not only on T cells but also on pituitary cells in both humans and mice (27). In mouse models injected with ipilimumab, complement deposition and lymphocyte infiltration have been observed in the pituitary gland, suggesting that anti-CTLA-4 antibodies directly interact with CTLA-4 expressed on pituitary cells, leading to immune-mediated damage (27). We observed that the clinical features of CPHD resemble those of hypophysitis, particularly headaches and pituitary swelling. These manifestations may reflect the direct impact of anti-CTLA-4 antibodies on the pituitary gland. Additionally, given that a substantial proportion of CPHD cases were accompanied by non-endocrine irAEs, it is plausible that both CPHD and these irAEs represent a common disease entity characterized by direct immune-mediated injury driven by anti-CTLA-4 antibodies.

Regarding the molecular mechanisms of IAD, a previous study identified corticotroph-specific antibodies in 10% of patients with pituitary irAEs, all of whom had tumors exhibiting ectopic ACTH expression (28). The authors proposed that ectopic proopiomelanocortin expression in tumors triggers the activation of autoreactive T cells, which is further amplified by ICIs, leading to selective corticotroph injury and ACTH deficiency (29). However, substantial number of IAD cases without ectopic ACTH expression have also been reported (28). In our study, the increased frequency of IAD in the Nivo/Ipi group suggests a potential synergistic effect between anti-PD-1 and anti-CTLA-4 antibodies in the development of IAD. PD-1 and CTLA-4 blockade enhance immune responses through distinct mechanisms: PD-1 blockade primarily expands tumor-infiltrating CD8^+^ T cells, whereas CTLA-4 blockade promotes the activation of both CD8^+^ T cells and CD4^+^ effector T cells (30). It is suggested that CD8^+^ T cells may be involved in the development of IAD, while CD4^+^ effector T cells may play a role in CPHD. To confirm this hypothesis, further investigation from the perspective of immune checkpoint biology is needed.

The limitations of this study are listed as follows. Although we observed a high incidence of pituitary irAEs in this cohort, some cases lacked results of laboratory test and MRI, which may have led to an underestimation of the true incidence, particularly of CPHD. As reported in our previous study, ACTH, cortisol, and thyroid function were assessed every 4 weeks during nivolumab monotherapy at our institution, with high compliance rate (14). Given that the frequent monitoring was also applied during combination immunotherapy, most cases were likely detected. Some patients in the Nivo/Ipi group had prior exposure to ICIs, which may have influenced the frequency or timing of irAE onset. We attempted to minimize this potential bias: there was no apparent difference in irAE frequency (Supplementary Table 1). Finally, as this study was conducted at a single center, further validation is warranted.

Based on the results of this study, we propose a perspective on managing pituitary irAEs in the context of combination immunotherapy. Compared to PD-1 monotherapy, combination immunotherapy is associated with a significantly higher incidence of pituitary irAEs, which may develop very early, even after the first course of treatment. Therefore, it may be advisable for physicians to regularly monitor cortisol levels to facilitate early detection of pituitary irAEs. On the other hand, CPHD and IAD are distinct clinical entities that require different management strategies. CPHD may require higher doses of glucocorticoids than IAD and relapse following re-administration of Nivo/Ipi. Patients should be appropriately informed of this risk in advance. Recognizing these differences between CPHD and IAD is essential for the optimal management of pituitary irAEs during combination immunotherapy.

## Data Availability

All data produced in the present study are available upon reasonable request to the authors

## Acknowledgments

This work was supported by Japan Society for the Promotion of Science KAKENHI Grant Number 19K23942 and 25K19669.

## Author Contributions

AY and TH collected and analyzed the data. IY analyzed the data and provided funding. KM, MN, SK, DK, TS, HF, KO, YU, and DT contributed to the discussion. DY supervised the entire study. AY, TH and IY drafted the manuscript, and all authors reviewed, edited, and approved the manuscript.

## Disclosures

IY received lecture fees from Ono Pharmaceutical Co., Ltd., Bristol-Myers Squibb, and MSD KK. DY has received consulting/lecture fees from Eli Lilly Japan K.K., Kyowa Kirin Co., Ltd., Nippon Boehringer Ingelheim Co., Ltd., Novo Nordisk Pharma Ltd, Sanofi K.K, and Sumitomo Pharma Co., Ltd.; and research funding/grants from Arkray Inc., the Japan Association for Diabetes Education and Care, Nippon Boehringer Ingelheim Co., Ltd., Novo Nordisk Pharma Ltd., Taisho Pharmaceutical Co., Ltd., and Terumo Corporation.

## Data Availability

The data sets analyzed during the current study are not publicly available but are available from the corresponding author on reasonable request.

## Notes

### Competing Interest Statement

The authors have declared no competing interest.

### Author Declarations

Ethics committee/IRB of the Kyoto University Graduate School of Medicine gave ethical approval for this work.

