## Supplemental Data 1 for "Distinct Patterns of Pituitary Dysfunction in Combination Immunotherapy with Nivolumab and Ipilimumab"

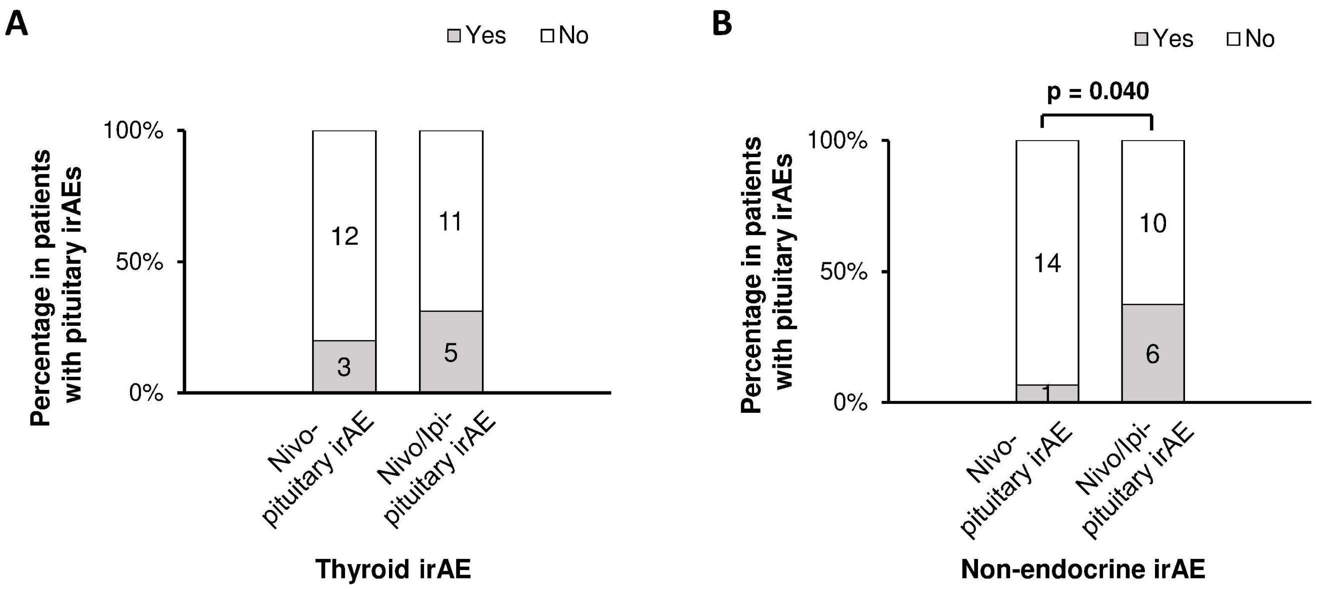


**Supplementary Figure 1. Incidence of other irAEs in patients with pituitary irAEs according to ipilimumab use.** irAE, immune-related adverse event; Nivo, nivolumab monotherapy; Nivo/Ipi, combination therapy using nivolumab and ipilimumab. (A) Incidence of thyroid irAEs, (B) Incidence of non-endocrine irAEs. Statistical analyses were performed using the Pearson's chi-square test. Significant differences are shown in bold.

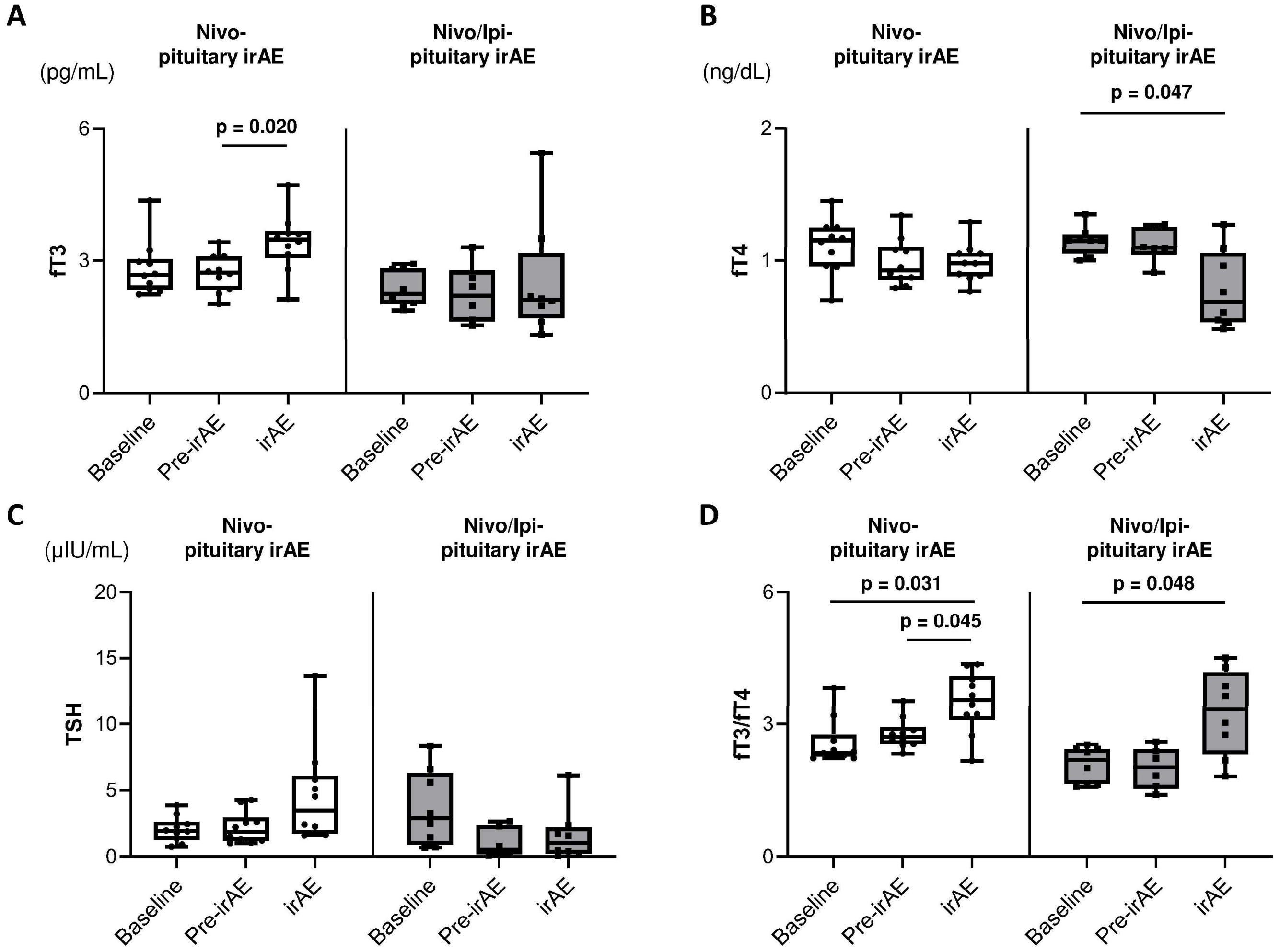


**Supplementary Figure 2. Differences in thyroid function among patients with pituitary irAEs according to ipilimumab use.** Longitudinal data at three time points, baseline, the last measurement before pituitary irAE (pre-irAE), and at pituitary irAE diagnosis (irAE): free T3 (fT3) (A), free T4 (fT4) (B), TSH (C), and fT3/fT4 ratio (D). Statistical analyses were made using the Steel-Dwass test. We express significant differences of $p < 0.05$ in boldface text and do not show results for $p \geq 0.05$.

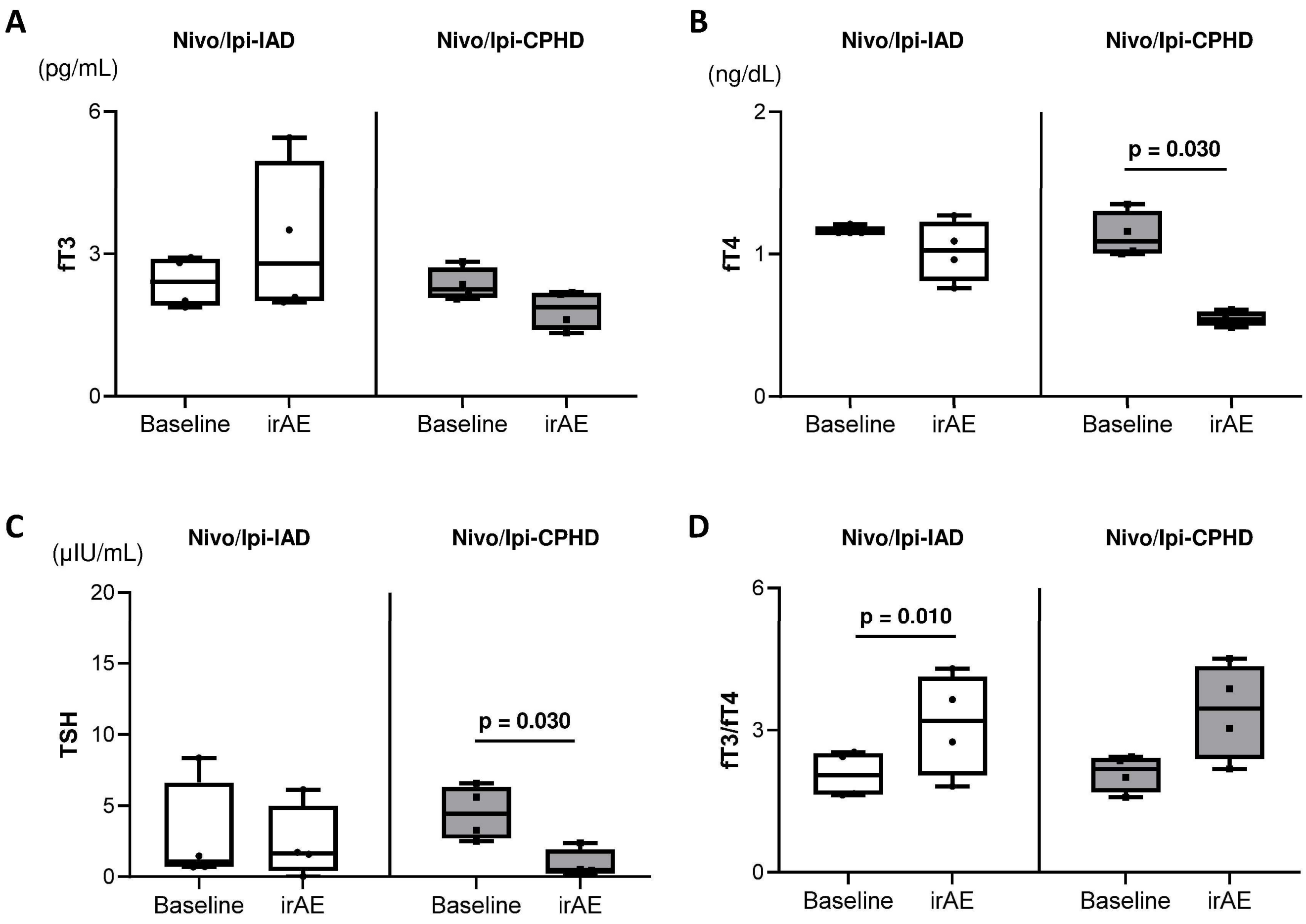


**Supplementary Figure 3. Differences in thyroid function among patients with pituitary irAEs induced by Nivo/Ipi therapy according to the patterns of pituitary dysfunction.** IAD, isolated ACTH deficiency; CPHD, combined pituitary hormone deficiency. fT3 (A), fT4 (B), TSH (C), and fT3/fT4 ratio (D). Statistical analysis was performed using the Mann-Whitney U test. We express significant differences of $p < 0.05$ in boldface text, and do not show results for $p \geq 0.05$.

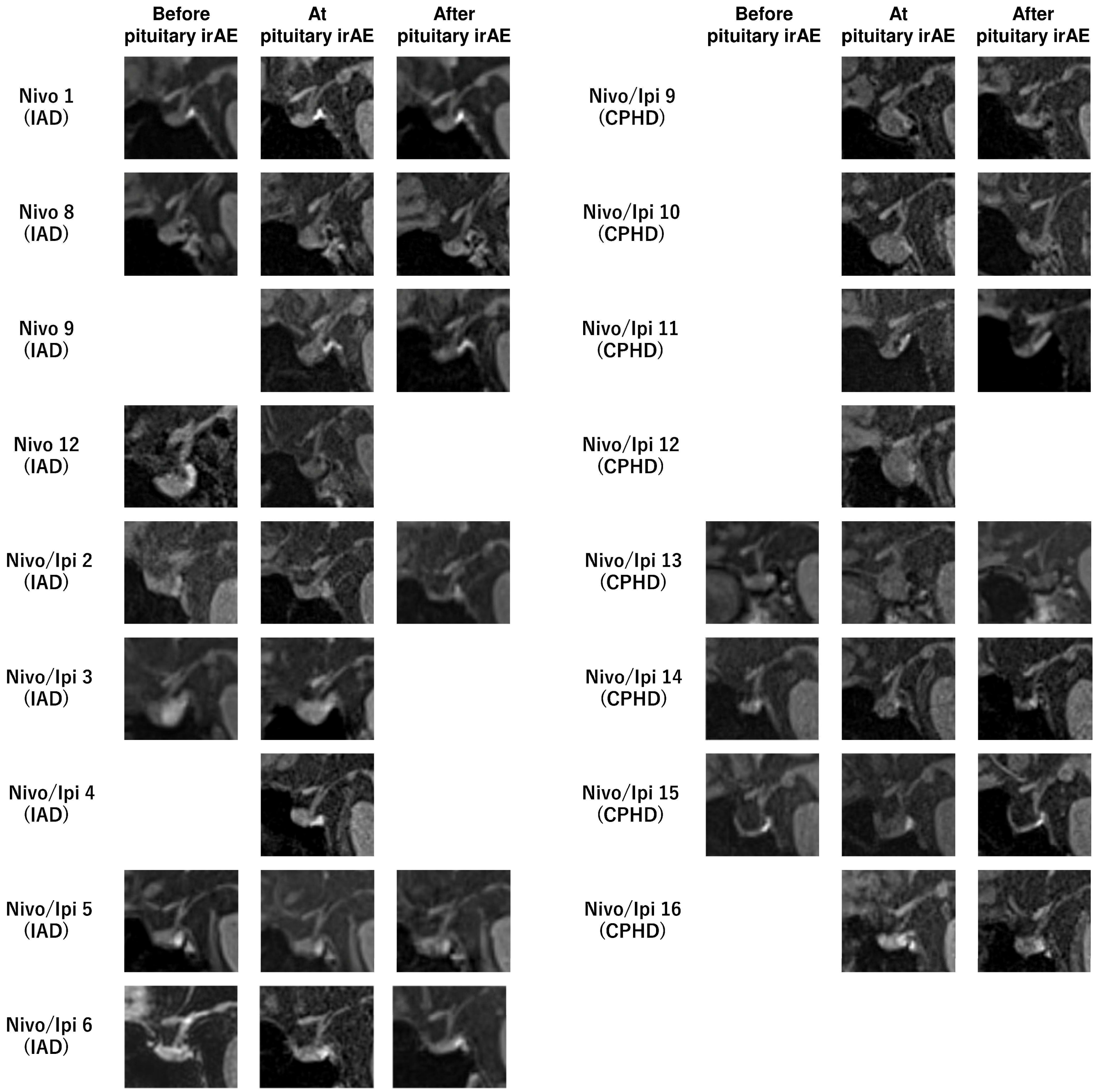


**Supplementary Figure 4. Pituitary MRI results in patients with pituitary irAEs.** Pituitary magnetic resonance imaging (MRI) results obtained at three time points: before the onset of pituitary irAE (before pituitary irAE), at the time of pituitary irAE diagnosis (at pituitary irAE), and after the onset of pituitary irAE (after pituitary irAE). Time points for after pituitary irAE are as follows; Nivo 1, day 83; Nivo 8, day 554; Nivo 9, day 169; Nivo/Ipi 2, day 116; Nivo/Ipi 5, day 148; Nivo/Ipi 6, day164; Nivo/Ipi 9, day 61; Nivo/Ipi 10, day 45; Nivo/Ipi 11, day 153; Nivo/Ipi 13, day 531; Nivo/Ipi 14, day 190; Nivo/Ipi 15, day 103; Nivo/Ipi 16, day 33.

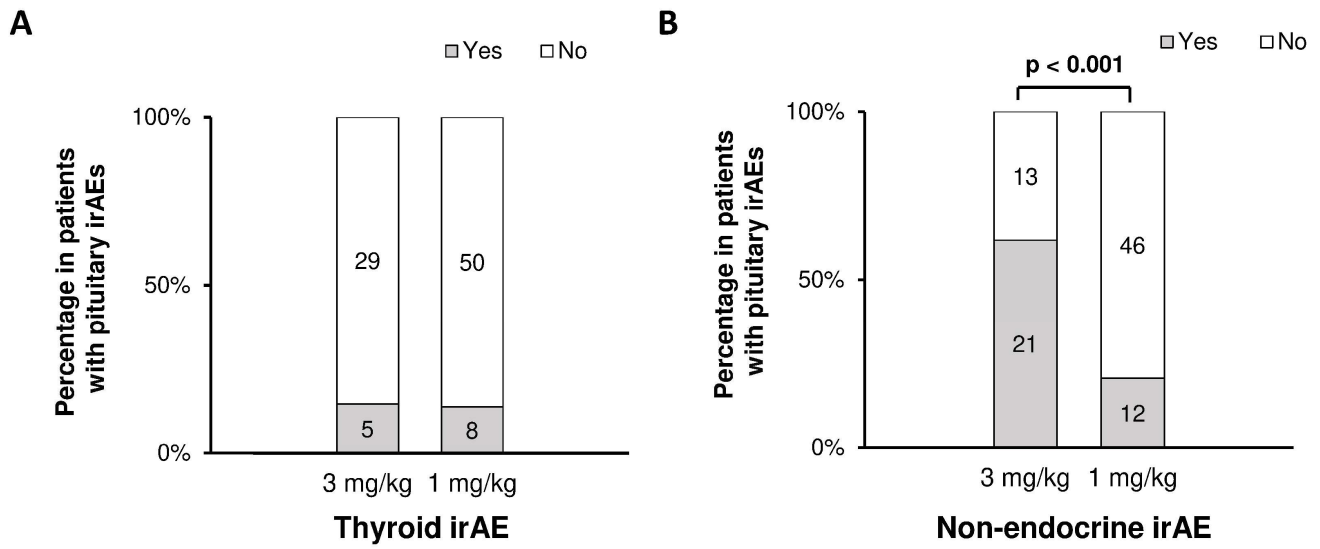


**Supplementary Figure 5. Incidence of other irAEs in the Nivo/Ipi group according to dosage of ipilimumab.** Incidence of thyroid irAEs (A) and non-endocrine irAEs (B). Statistical analyses were performed using the Pearson's chi-square test. Significant differences are shown in bold.
