## Supplementary Table 1 for "Distinct Patterns of Pituitary Dysfunction in Combination Immunotherapy with Nivolumab and Ipilimumab"

**Supplementary Table 1. Comparison of patient characteristics according to prior ICI use in the Nivo/Ipi group**

|  | **Prior ICI (+)**  **(n = 31)** | **Prior ICI (−)**  **(n = 61)** | ***p*** |
| --- | --- | --- | --- |
| Age (year) | 71 (56–73) | 69 (56–73) | 0.182 |
| Gender (n) |  |  | 0.662 |
| Male | 21 (66.5%) | 44 (70.7%) |  |
| Female | 10 (33.5%) | 17 (29.3%) |  |
| Primary sites (n) |  |  | **< 0.001** |
| Malignant melanoma | 21 (67.7%) | 13 (21.3%) |  |
| Lung cancer | 8 (25.8%) | 25 (41.0%) |  |
| Others | 2 (6.5%) | 23 (37.7%) |  |
| Renal cell carcinoma | 1 (3.2%) | 12 (19.7%) |  |
| Esophageal cancer | 0 (0.0%) | 7 (11.5%) |  |
| Malignant pleural mesothelioma | 1 (3.2%) | 4 (6.6%) |  |
| irAE (n) | 9 (29.0%) | 18 (29.5%) | 0.962 |
| Pituitary irAE | 6 (19.4%) | 10 (16.4%) | 0.723 |
| Thyroid irAE | 3 (9.7%) | 10 (16.4%) | 0.382 |
| Diabetes mellitus | 2 (6.5%) | 1 (1.6%) | 0.219 |
| Non-endocrine irAE (n) | 15 (48.4%) | 18 (29.5%) | 0.074 |
| Pneumonitis | 3 (9.7%) | 3 (4.9%) |  |
| Dermatopathy | 3 (9.7%) | 1 (1.6%) |  |
| Colitis | 2 (6.5%) | 3 (4.9%) |  |
| Hepatitis | 5 (16.1%) | 5 (8.2%) |  |
| Acute kidney injury | 1 (3.2%) | 1 (1.6%) |  |
| Others | 6 (19.4%) | 6 (9.8%) |  |

ICI, immune checkpoint inhibitor; Nivo/Ipi, combination therapy using nivolumab and ipilimumab; n, number of subjects; irAE, immune-related adverse event. ‘Others’ in non-endocrine irAE includes the following events: fever (n = 2), pancreatitis (n = 1), esophagitis (n = 1), peripheral neuropathy (n = 1), and laryngitis (n = 1) in the Prior ICI (+) group: myocarditis (n = 2), fever (n = 1), esophagitis (n = 1), polymyalgia rheumatica (n = 1), and myositis (n = 1) in the Prior ICI (−) group. Data of continuous variables are expressed as medians (interquartile range). Statistical analyses were performed using the Mann-Whitney U test for continuous variables and the Pearson’s chi-square test for categorical variables. Significant differences are shown in bold.
