## Supplementary Table 4 for "Distinct Patterns of Pituitary Dysfunction in Combination Immunotherapy with Nivolumab and Ipilimumab"

**Supplementary Table 4. Comparison of patient characteristics according to dosage of ipilimumab in the Nivo/Ipi group**

|  | **3 mg/kg (n = 34)** | **1 mg/kg (n = 58)** | ***p*** |
| --- | --- | --- | --- |
| Age (year) | 70 (58–74) | 69 (56–74) | 0.607 |
| Gender (n) |  |  | 0.235 |
| Male | 23 (67.7%) | 42 (72.4%) |  |
| Female | 11 (32.3%) | 16 (27.6%) |  |
| Primary site (n) |  |  | **< 0.001** |
| Malignant melanoma | 34 (100.0%) | 0 (0.0%) |  |
| Lung cancer | 0 (0.0%) | 33 (56.9%) |  |
| Others | 0 (0.0%) | 25 (43.1%) |  |
| Renal cell carcinoma | 0 (0.0%) | 13 (22.4%) |  |
| Esophageal cancer | 0 (0.0%) | 7 (12.1%) |  |
| Malignant pleural mesothelioma | 0 (0.0%) | 5 (8.6%) |  |
| irAE (n) |  |  |  |
| Pituitary irAE | 9 (26.5%) | 7 (12.1%) | 0.079 |
| Thyroid irAE | 5 (14.7%) | 8 (13.8%) | 0.903 |
| Diabetes mellitus | 2 (5.9%) | 1 (1.7%) | 0.278 |
| Non endocrine-related irAE (n) | **21 (61.8%)** | **12 (20.7%)** | **<0.001** |
| Pneumonitis | 3 (8.8%) | 3 (5.2%) |  |
| Dermatopathy | 4 (11.8%) | 0 (0.0%) |  |
| Colitis | 5 (14.7%) | 0 (0.0%) |  |
| Hepatitis | 9 (26.5%) | 1 (1.7%) |  |
| Acute kidney injury | 1 (2.9%) | 1 (1.7%) |  |
| Others | 5 (14.7%) | 7(12.1%) |  |

Ipilimumab is administered every 3 weeks for malignant melanoma and renal cell carcinoma, and every 6 weeks for lung cancer, esophageal cancer, and malignant mesothelioma. ‘Others’ in non-endocrine-related irAE includes the following events: fever (n = 2), esophagitis (n = 1), pancreatitis (n = 1), and laryngitis (n = 1) in the 3 mg/kg ipilimumab group; polymyalgia rheumatica (n = 1), esophagitis (n = 1), peripheral neuropathy (n = 1), myocarditis (n = 1), fever (n = 1), and myositis (n = 2) in the 1 mg/kg ipilimumab group. Data of continuous variables are expressed as medians (interquartile range). Statistical analyses were performed using the Mann-Whitney U test for continuous variables and the Pearson’s chi-square test for categorical variables. Significant differences are shown in bold.
